# Carbapenem-resistant *Klebsiella pneumoniae* in university-affiliated hospitals: risk factors for isolation among hospitalized patients and molecular subtyping

**DOI:** 10.1101/2022.02.08.22269570

**Authors:** Ketevan Kobaidze, Jesse Jacob, Traci Leong, W. Dana Flanders

## Abstract

**Background:** Carbapenem-resistant *Klebsiella pneumoniae* (CRKP) is an important healthcare-associated pathogen. This study aimed to identify factors associated with CRKP isolation among hospitalized patients, describe molecular epidemiology, and mortality associated with CRKP isolation.

**Methods:** We performed a retrospective case-control study at the two university-affiliated teaching hospitals. 150 patients were included (30 cases and 120 controls) in this study.

Each patient with CRKP, a case-patient, was matched with four controls by admission facility, age, and sex. Controls, patients without CRKP were randomly selected from a computerized list of inpatients whose admission date was the same as that of the case, within 48 hours of the date of the initial positive culture. We calculated the risk of in-hospital death as the number of deaths divided by the number of cases and evaluated the risk of mortality associated with the site of positive culture. Molecular epidemiology investigation using comparison of restricted DNA patterns of CRKP by pulsed-field gel electrophoresis (PFGE) was conducted.

**Results:** A greater proportion of cases than controls had undergone an invasive procedure, including use of a central vein catheter (p=0.007, OR, 3.4, 95% CI, 1.4-8.7), and mechanical ventilation (p=0.002, OR, 3.6, 95% CI, 1.6-8.1), nutrition by tube feeding (p=0.001, OR, 4.2, 95% CI, 1.8 −10).

Pre-admission treatment within two months with the following antibiotic classes was associated with CRKP isolation: carbapenems (p=0.001, OR, 24.4, 95% CI, 2.73-217.96), fluoroquinolones (p<0.0001, OR, 6.17, 95% CI, 2.4 – 15.83), anti-pseudomonal penicillin (p = 0.02; OR, 6.03; 95% CI, 1.98 −18.32), and cephalosporins (p=0.001, OR, 5.36, 95% CI, 2.07 −13.87).

The molecular analysis detected that over 90% of isolates shared similar PFGE patterns.

CRKP isolation was associated with significantly higher In-hospital mortality (36.7% vs 3.3%) in comparison to controls (p<.0001).

Positive cultures from sites other than urine were associated with substantially higher mortality than was a positive urine culture (RR= 4.0).

## Introduction

For the past decade, national surveillance studies have documented a growing rate of antimicrobial resistance in hospitals in the United States. ^1^ Infection caused by multidrug-resistant Gram-negative bacteria present a challenge to clinicians since it is associated with increased mortality, morbidity, length of hospital stay, and financial cost. ^2^ The mode of acquisition and spread of resistant Gram-negative bacilli is complex. These organisms are highly efficient at acquiring multidrug-resistant coding genes under selective antibiotic pressure and are capable of rapid dissemination. ^3, 4^

The global rise in antimicrobial resistance coupled with the lack of new anti-microbial agents increasingly drives treatment choices toward carbapenems. Carbapenems, the most enzyme-stable class of β-lactam antibiotics, have been considered the last resort antimicrobial therapy against many antibiotic-resistant Gram-negative organisms.

Over the past decade, carbapenem-resistant *Klebsiella pneumoniae* (CRKP) has emerged as a serious healthcare-associated pathogen in the United States and worldwide. ^3, 4, 5, 6^ CRKP threatens hospitalized patients and lengthens recovery since very limited or no drugs are available to treat this infection. Large city-wide outbreaks have been reported in northeastern states, where it became endemic.

The first CRKP isolate in the Emory Healthcare system was detected in September 2006. The patient was from New York City, an endemic area for CRKP, seeking evaluation for a wound infection following liver transplantation. Because there were frequent transfers within the Emory Healthcare system (two 500-bed acute care hospitals, 1 long-term acute care facility (LTAC), 2 rehabilitation units, and 1 geriatric hospital), there was a great potential for spread.

We conducted a case-control study to examine factors associated with isolation of CRKP among the hospitalized population in the two acute care hospitals, Emory University Hospital Midtown (EUHM) and Emory University Hospital (EUH).

## Methods

All adult patients aged≥18 years who had been treated in the hospital from September 2006 through December 2008 and had a clinical culture positive for CRKP were enrolled. A case-the patient was identified after the initial CRKP culture was reported, and included only once, regardless if CRKP was isolated further.

Each case-patient was matched with four control-patients by admission facility, age +/-5 years, sex, and date of culture, as a time-at-risk for hospitalized patients.

Controls, patients without CRKP were randomly selected from a computerized list of inpatients who matched the case age (+/-5 years), sex, and facility and whose admission date was within 48 hours of the date of the initial, positive culture. Patients, who spent less than 48 hours in the hospital, and obstetric service patients were not included as controls. We chose the control group from the population at risk which allowed us to estimate the risk associated with specific exposures, such as prior use of antimicrobial agents. ^7^ It also allowed us to compare the mortality associated with CRKP isolation to that without CRKP. A health-care exposure was defined as hospitalization of more than 48h, previous hospitalization within six months, previous LTAC or nursing home placement, or hemodialysis.

From the medical record, we abstracted information including basic demographics, admission date, outcome, services, and admission facilities, urinary catheter use, central line placement, intubation, and mechanical ventilation before CRKP isolation, antibiotic use within two months before hospitalization as it was documented in the chart, or electronic records and in the hospital prior CRKP isolation; exposure to interventional procedures, tube feeding, and length of stay in the hospital. The performance status was measured by the Karnofsky scale per physical and occupational services evaluation at the time of CRKP isolation.

The Charlson comorbidity scores were used to assess and compare patients’ co-morbidities, ^8^ and APACHE-II scores were compared for those who were treated in the ICU.

Laboratory confirmation of CRKP was restricted to detection of *bla-KPC* gene by PCR. ^9^ For PCR, the bacterial DNA was extracted using the EZ-1 BioRobot (Qiagen, Germantown, MD). Amplification and detection of a 399 base pair segment of the *bla-KPC* gene were performed on the 7500 Real-Time PCR system (Applied Biosystems, Foster City, CA). All isolates were found to contain the *bla*-KPC gene by PCR. Molecular subtyping by PFGE was performed to detect genetic relatedness among isolates.

Analyses. Descriptive analyses were conducted, including means, medians, and percentiles. The comparison of clinical characteristics between cases and controls was made using Chi-Square (or Fisher’s exact) tests. Relative risks (RR), and exact confidence intervals (CI) for the association between risk factors and CRKP were estimated using exact conditional logistic regression (Proc Exact, in SAS, version v9.2). Conditional logistic regression was used due to the large degrees of freedom in the model from our matched design. Exact distributions were used due to the small numbers for some combinations of exposure and disease. In separate analyses of the cases, we calculated the risk of in-hospital death as the number of deaths divided by the number of cases. To assess the association of mortality with the site of culture, we calculated the risk ratio: the proportion who died among those for whom CRKP was isolated from a site other than urine divided by the proportion who died among those for whom CRKP was isolated from the urine.

Molecular subtyping by pulsed-field gel electrophoresis (PFGE) was performed for the comparison of restricted DNA patterns of all CRKP isolates. ^10^ For better molecular characterization additional 21 isolates from all sites (total 51 isolates) were included in molecular subtyping including our 30 isolates.

This study was approved by the Institutional Review Board of the Emory University School of Medicine.

## Results

### Study population

The first case was identified in September of 2006, 13 cases were detected in 2007, and 16 cases in 2008 across these two hospitals (total of 30 with 120 matched controls). No seasonal variations were apparent in the identification of CRKP.

The clinical characteristics of patients with CRKP enrolled in this study (summarized in Table 1), show the median age was 60 years. Urine was the most common site of organism isolation, followed by lung and wound. Of 30 cases eleven patients died (36.7%) whereas 4 patients in the control group (3.3%) died. Nine of the 11 deceased cases had CRKP isolated from specimens other than urine or multiple sites. Cases with a positive culture from a site other than urine had substantially higher mortality than patients whose positive cultures were only from urine (RR= 4.0). Four patients had received solid organ transplants before CRKP isolation, and two of those died in the hospital.

**Table 1.**
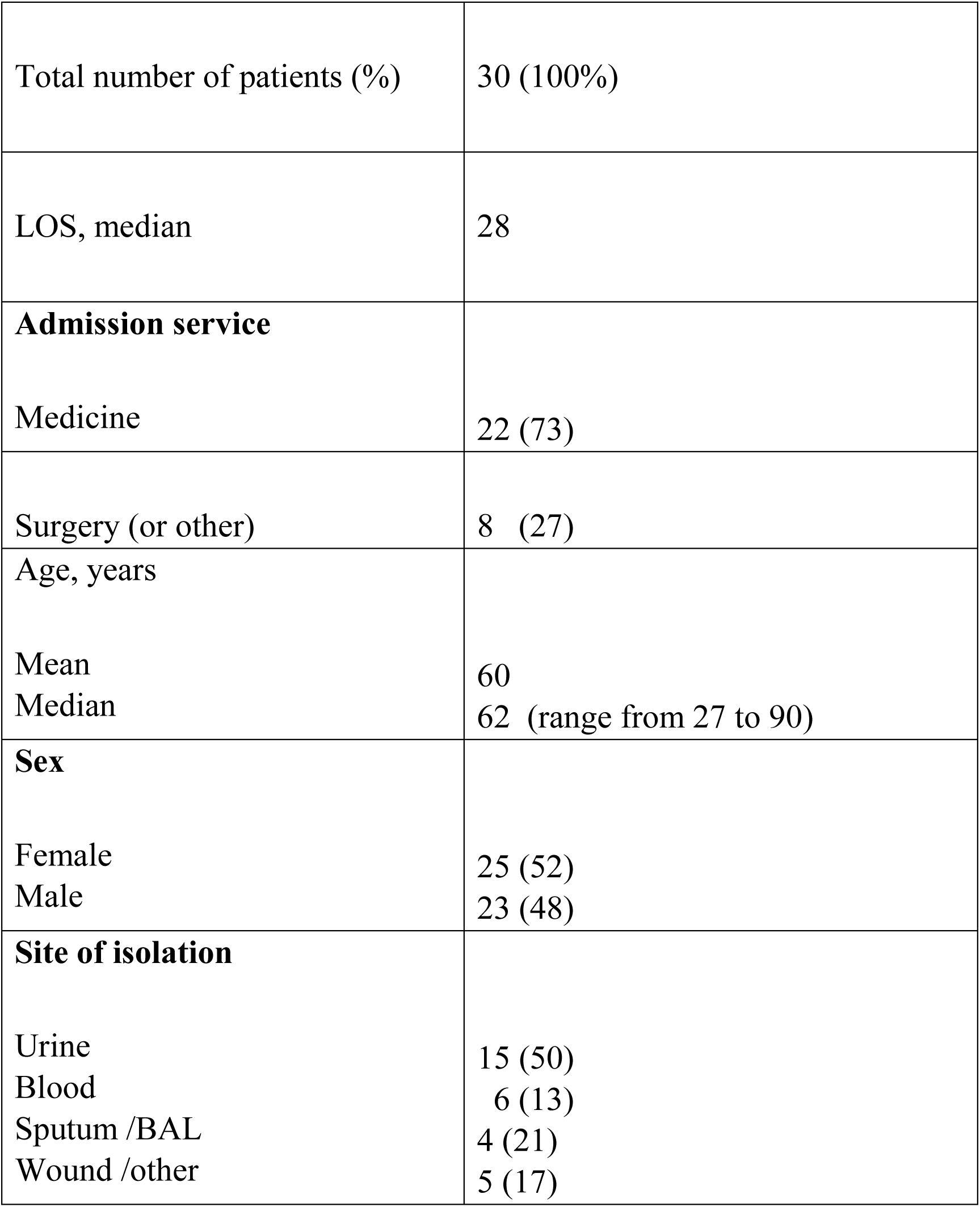
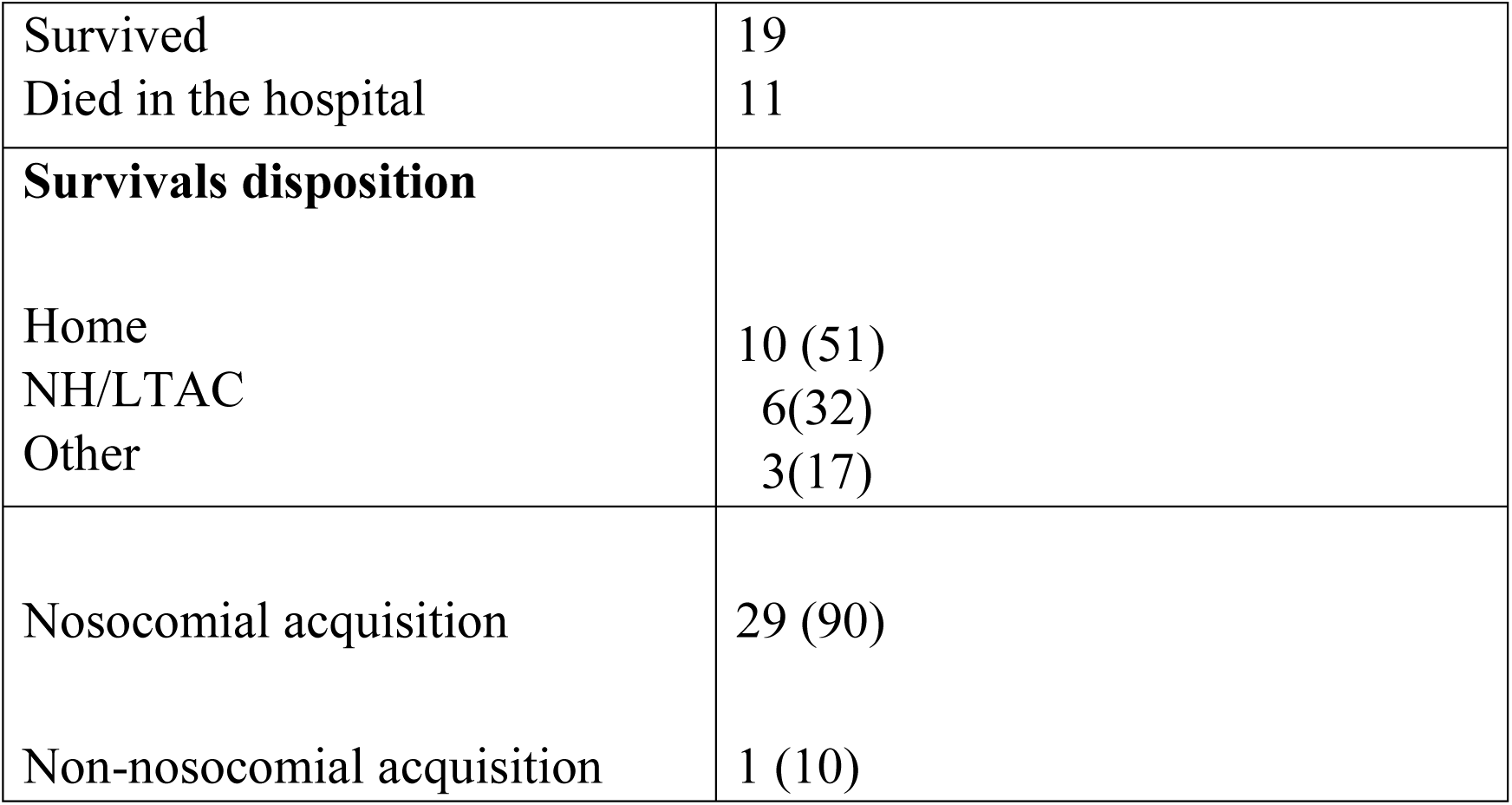
Case-patients characteristics.

|  |  |
| --- | --- |
| Total number of patients (%) | 30 (100%) |
| LOS, median | 28 |
| **Admission service** |  |
| Medicine | 22 (73) |
| Surgery (or other) | 8 (27) |
| Age, years |  |
| Mean | 60 |
| Median | 62 (range from 27 to 90) |
| **Sex** |  |
| Female | 25 (52) |
| Male | 23 (48) |
| **Site of isolation** |  |
| Urine | 15 (50) |
| Blood | 6 (13) |
| Sputum /BAL | 4 (21) |
| Wound /other | 5 (17) |
| Survived | 19 |
| Died in the hospital | 11 |
| **Survivals disposition** |  |
| Home | 10 (51) |
| NH/LTAC | 6(32) |
| Other | 3(17) |
| Nosocomial acquisition | 29 (90) |
| Non-nosocomial acquisition | 1 (10) |

All but one patient was confirmed to have healthcare-acquired CRKP. The source of CRKP acquisition in one patient was not identified and was considered to be non-healthcare related

### Risk factors associated with isolation of CRKP

Factors assessed for association with isolation of CRKP in the hospital are summarized in Table 2. A greater proportion of cases than controls had undergone an invasive procedure, including the use of a central vein catheter (p=0.007, OR, 3.4, 95% CI, 1.4-8.7), and mechanical ventilation (p=0.002, OR, 3.6, 95% CI, 1.6-8.1). Nutrition by tube feeding (p=0.001, OR, 4.2, 95% CI, 1.8 − 10) and admission from a nursing home or LTAC were also associated with increased risk (p<0.001, OR=6.2, 95% CI, 2.5-15.1).

**Table 2.** Risk factors associated with acquisition of KPC-KP among hospitalized patients: univariate analysis of risk factors associated with acquisition of KPC-KP

|  | Case | Control | P-Value | Odds Ratio | Confidence Interval |
| --- | --- | --- | --- | --- | --- |
| **Antibiotics (pre hospital)** |  |  | <.001 |  |  |
| 0 | 11<br>(36.7%) | 100 (81.3%) |  |  |  |
| 1 | 4 (13.3%) | 7 (5.7%) | .493 | 1.639 | (.39,6.76) |
| 2+ | 15 (50%) | 16 (13%) | <.001 | 8.547 | (3.3,21.74) |
| **antipseudomonal PCN** | 8 (26.7%) | 7 (5.7%) | .002 | 6.026 | (1.98,18.32) |
| **Cephalosporines** | 11<br>(36.7%) | 12 (9.8%) | .001 | 5.355 | (2.07,13.87) |
| **Aminoglycosides** | 3 (10.0%) | 2 (1.6%) | .042 | 6.722 | (1.07,42.21) |
| **Vancomycin** | 2 (6.7%) | 9 (7.3%) | 1.0 | .905 | (.19,4.42) |
| **Levofloxacin/quinolones** | 12 (40%) | 12 (9.8%) | <.0001 | 6.167 | (2.40,15.83) |
| **carbapenem** | 5 (16.7%) | 1 (.85) | .001 | 24.4 | (2.73,217.96) |
| **Antibiotics (during hospital)** |  |  | .005 |  |  |
| 0 | 1 (3.3%) | 18 (14.6%) |  |  |  |
| 1 | 5 (16.7%) | 37 (30.1%) | 0.99 | 0 | (1.29, 10.64) |
| 2+ | 24 (80%) | 48 (39%) | .015 | 3.7 | (1.78, 29.31) |
| **Carbapenem** | 15 (50%) | 4 (3.3%) | .016 | 12.5 | (1.59,100) |
| **Cephalosporines** | 4 (13.3%) | 39 (31.7%) | .068 | .331 | (.11,1.02) |
| **Aminoglycosides** | 2 (6.7%) | 6 (4.9%) | .655 | 1.393 | (.27,7.27) |
| **antipseudomonal PCN** | 20<br>(66.7%) | 29 (23.6%) | <.001 | 6.483 | (2.73,15.41) |
| **Vancomycin** | 5 (16.7%) | 16 (13.0%) | .565 | 1.34 | (.45, 3.99) |
| **Levofloxacin/quinolones** | 16<br>(53.3%) | 38 (30.9%) | .032 | 2.556 | (1.13,5.76) |
| **Intubated/Vent** | 16<br>(53.3%) | 30 (24.4%) | 0.002 | 3.55 | (1.55, 8.13) |
| **Length of Stay** |  |  | .001 | 1.029 | (1.01,1.05) |
| **Charlson Score** |  |  | 0.047 | 1.123 | (1.00,1.26) |
| **Urinary Catheter** | 24 (80%) | 85 (69.1%) | .270 | 1.79 | (0.68, 4.74) |
| **APACHE II for ICU** |  |  | .058 | 1.036 | (.99,1.07) |
| **Central Vein Catheter** | 23<br>(76.7%) | 60 (48.8%) | .007 | 3.44 | (1.38, 8.62) |
| **Tube Feeding** | 14<br>(46.7%) | 21 (17.1%) | .001 | 4.26 | (1.80, 10) |
| **Pre-hospital Site** |  |  |  |  |  |
| NH + LTAC + Hospital | 22<br>(73.3%) | 38 (30.9%) | <.001 | 6.151 | (2.51, 15.06) |
| Home | 8 (26.7%) | 85 (69.1%) |  |  |  |
| **KPS score** |  |  | <.001 | 0.934 | (0.91, 0.96) |

Most patients with CRKP were treated with two or more antibiotics within 2 months prior to current hospitalization (p<0.005, OR, 8.5, 95% CI, 3.3 −21.7). Pre-admission treatment two months before hospitalization with the following antibiotic classes was associated with CRKP acquisition: carbapenems (p=0.001, OR, 24.4, 95% CI, 2.73-217.96), fluoroquinolones (p<0.001, OR, 6.17, 95% CI, 2.4 – 15.83), anti-pseudomonal penicillin (p<0.02, OR, 6.03, 95% CI, 1.98 −18.32), and cephalosporins (p = 0.001, OR, 5.36, 95% CI, 2.07 −13.87).

Post-admission treatment with anti-pseudomonal penicillins (p<0.001, OR, 6.5, 95% CI, 2.7-15.4), carbapenems (p=0.016, OR, 12.4, 95% CI, 1.59 −100), and a fluoroquinolone (p<0.03, OR, 2.6, 95% CI, 1.1-5.7) were significantly associated with CRKP isolation. Interestingly cephalosporin antibiotic use in the hospital was not a significant risk factor (Table 2).

On average cases stayed in the hospital 32.5 days compared to 10 days for controls, and the average time from admission to first CRKP isolation was 12.3 days. A Charlson score of seven or higher (p=0.047, OR, 1.12, 95% CI, 1.0 – 1.26), and low scores of performance status by Karnofsky scale (p<0.001, OR, 0.93, 95% CI, 0.91-0.96) were significantly associated with the isolation of CRKP.

### Molecular subtyping

The PFGE pattern analysis (Fig 1) showed that over 90% of the isolates shared similar PFGE patterns (>85% band similarity). In addition, the prevailing pattern was found to be very similar to PFGE patterns of isolates that CDC determined to be multilocus sequential type ST258, and which appears to be the dominant CRKP strain seen throughout the United States. ^11^

**Figure 1.**
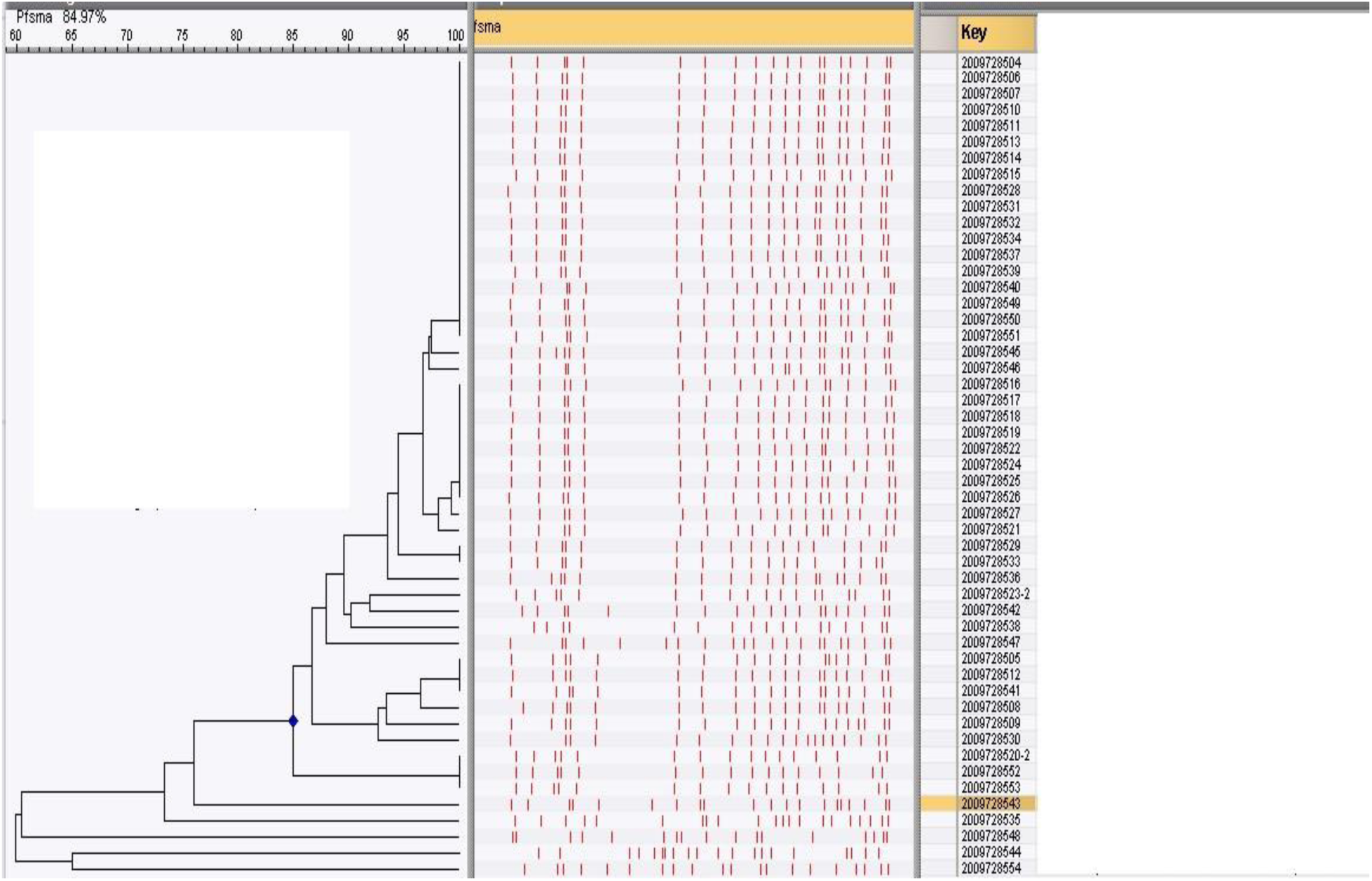
Dendrogram of similarity of PFGE patterns. Isolates above #2009728543 has 85% similar band patterns, 5 isolates pattern has distinct patterns.

## Discussion

Since its isolation in 1996 in North Carolina CRKP has been detected in multiple states ^12^ and across the world including China, ^13^ Europe, ^3, 14^ Central and South America ^15, 16^ and Israel. ^17^ It is a challenging infection with attributable mortality as high as 40 %, ^18^ and it is rapidly spreading. CRKP is endemic in the Northeastern United States, particularly in the New York area, but periodic outbreaks have been reported throughout the country. ^19, 20, 21^ Some authors speculate that the global spread could be greater than reported since the detection of CRKP by clinical microbiology laboratories remains difficult.

Previous studies have identified the inter-hospital spread of CRKP ^18, 19, 20^ and the results of our study also suggest that CRKP was introduced in our system by an index case in September 2006, and subsequently spread through these hospitals. Molecular sub-typing by PFGE confirmed >85% genetic relationship among the isolates. Our results are consistent with previous reports that CRKP dissemination occurs in part by the spread of isolates from patients previously hospitalized in an endemic area. ^11, 22^ Two of our patients traveled from New York City and sought medical attention in our facilities.

Even though the number of incidents remained low we observed a significant increase in incidence during the study period. Detection of CRKP infection likely represents only part of the problem since the vast majority of transmissions likely occur through colonized asymptomatic patients. Infection spread was well documented in this study, showing how a strain of CRKP can quickly disseminate throughout a hospital

Most prior studies of risk factors for CRKP acquisition have focused on antimicrobial exposure, underlying conditions, ICU exposure, and procedures. ^5, 6, 23, 24, 25, 26, 27^ The present study confirmed the use of multiple (two or more) antibiotics, including cephalosporins, fluoroquinolones, anti-pseudomonal penicillins, and carbapenems, before hospitalization was associated with increased risk isolation of CRKP. Previous research by Gasink and colleagues ^28^ had identified that fluoroquinolone use can be a risk factor for the acquisition of CRKP which is consistent with the results of our study.

The *bla-KPC* gene, responsible for resistance, resides on a transmissible plasmid and its relocation from other *Enterobacteriaceae* species has been elegantly confirmed by phage-typing. ^29^ Mathers, et al. have shown several mechanisms by which the CRKP resistance gene can spread including plasmid transfer and clonal spread.^23^ We found the occurrence of CRKP-producing *Klebsiella oxytoca* and *Citrobacter freundii* isolates in addition to CRKP in two of our cases. Regretfully we were not able to perform phage typing to confirm the resistance gene interspecies translocation.

Hussein and colleagues detected that exposure to at least one antibiotic drug before isolating CRKP was a risk factor.^20^ In contrast, our study identified that pre-hospital use of two or more antibiotics within two months significantly increased the risk of isolation of CRKP, whereas the use of one antibiotic before hospitalization didn’t reach a significant difference.

The results of the present study add to a substantial body of data demonstrating the role of previous antibiotic use in the development CRKP mechanism.

Our findings show that pre-hospital use of carbapenems was associated with a higher risk of acquiring CRKP. We also found that not only carbapenems but also fluoroquinolones and anti-pseudomonal antibiotic use were associated with significant risk while patients were treated in the hospital.

Interestingly, in-hospital cephalosporins use was not associated with a CRKP isolation risk, while within two months of pre-hospital use, this antibiotic was associated with a significantly increased risk of CRKP isolation.

Patients in our study had longer hospitalization, and underwent more interventional procedures including central line placement and intubation with mechanical ventilation, than controls. Case patients were treated in the ICU more often than controls (data not shown), APACHE II scores for those who were admitted to the ICU detected higher numbers of cases than controls but the difference was not statistically significant (Table 2).

Previous studies suggested that underlying multiple medical conditions were associated with a higher risk of acquisition of CRKP. ^26, 27, 28^ Our results support the conclusion that multiple comorbidities, here measured by the Charlson score, are a risk factor.

Hospitalized immunocompromised patients, including solid organ transplant recipients, are at increased risk to suffer from infection with multidrug-resistant pathogens. ^23^ Consistent with this finding is that four cases patients enrolled in our study were solid organ recipients, two of whom died in the hospital.

Although the increased risk for urinary tract infection and use of indwelling bladder catheter is well documented in CRKP by the authors, ^15, 30^ the study detected that most of our patients with this catheter had no greater risk for CRKP than without it. This finding was explained by the frequent use of indwelling catheters among all hospitalized patients.

CRKP outbreaks have also been reported in LTAC and nursing homes. ^30, 31, 32^ The present study identified admission from LTAC or nursing home as a risk factor associated with isolation of CRKP. This observation further supports the previous finding that CRKP is truly a healthcare-associated infection. Only one case-patient was admitted from their home, without identifiable exposure to healthcare. Although the number is small, it suggests that CRKP can be acquired in the community

Our study had several limitations. No surveillance screening was performed before this study to identify the CRKP carrier state initially. Due to the small number of cases, we decided to include all patients with CRKP, thus all patients documented to have infection or colonization were enrolled. Also due to the small sample size, we accommodated only for the matching factor, but not other covariates thus only univariate analysis was performed. The study was conducted in tertiary teaching hospitals close to LTAC, our finding cannot be generalized to all CRKP cases for other hospitals.

These findings have several important clinical implications. The use of multiple broad-spectrum antibiotics remains an important risk factor for developing CRKP. Moreover, hospitalized patients with multiple comorbidities and poor performance status are susceptible to CRKP, thus this risk group should be spared from unnecessary antibiotic exposure, and judicious use of antibiotics should be used by healthcare providers.

Consideration should be given when performing multiple invasive procedures since invasive procedures are associated with a higher risk for CRKP isolation.

CRKP infection poses a significant threat to the already vulnerable hospitalized patients. Efforts aimed at understanding and updating the epidemiology of CRKP among hospitalized patients should continue.

## Data Availability

All data produced in the present study are available upon reasonable request to the authors

